# Structural ethnic inequities in maternal mortality between Indigenous and non-Indigenous women in Paraguay, 2014–2023: a national analysis of territorial, institutional, and preventable factors

**DOI:** 10.64898/2026.06.22.26356286

**Authors:** Derlis Duarte-Zoilan, Juan Edgar Tullo Gomez, Beatriz Nuñez, Pasionaria Ramos

**Affiliations:** Department of Medicine, Autonomous University of Barcelona (UAB), Barcelona, Spain; Vall d’Hebron-Drassanes International Health Unit, Infectious Diseases Department, Vall d’Hebron University Hospital, Barcelona, Spain; Ministry of Public Health and Social Welfare (MSPYBS), General Directorate of Strategic Health Information (DIGIES), Asunción, Paraguay; Universidad del Norte, Asunción, Paraguay

**Keywords:** Maternal Mortality, Health Services, Indigenous, Healthcare Disparities, Maternal Health Services

## Abstract

**Background:** Indigenous women in Paraguay continue to experience disproportionately high maternal mortality despite national efforts to improve maternal health. Evidence on the structural factors underlying these disparities remains limited.

**Objectives:** To analyze structural ethnic inequities in maternal mortality between Indigenous and non-Indigenous women in Paraguay, focusing on territorial patterns, institutional access, and potentially preventable causes of death.

**Design:** National population-based study using maternal mortality records registered in Paraguay between 2014 and 2023. Maternal mortality ratios (MMRs), incidence rate ratios (IRRs), and absolute differences were estimated according to Indigenous status. Logistic regression models were used to assess associations with deaths occurring outside healthcare institutions and specific preventable causes of death.

**Results:** A total of 907 maternal deaths were identified, including 112 among Indigenous women (12.3%). Indigenous women were overrepresented by a factor of 4.8 relative to their population share. Maternal mortality remained consistently higher among Indigenous women throughout the study period, with mortality ratios ranging from 317.7 to 773.6 per 100,000 live births, compared with 58.7 to 145.1 among non-Indigenous women. Absolute inequalities remained persistently high over time. Overall, 24.3% of maternal deaths occurred outside healthcare institutions, with a substantially higher proportion among Indigenous women (44.6% versus 21.5%). After adjustment for age and educational level, Indigenous women had more than three times greater odds of dying outside healthcare institutions (aOR = 3.41; 95% CI: 2.20–5.29). Potentially preventable causes accounted for 42.4% of maternal deaths. Obstetric hemorrhage was strongly associated with Indigenous status (aOR = 3.83; 95% CI: 2.31–6.37).

**Conclusion:** Indigenous women in Paraguay experience a disproportionate burden of maternal mortality characterized by persistent ethnic disparities, higher occurrence of deaths outside healthcare institutions, and a substantial burden of preventable causes of death. These findings suggest the presence of enduring territorial, institutional, and healthcare access barriers that contribute to structural ethnic inequities in maternal health.

## Introduction

Maternal mortality remains one of the most sensitive indicators of inequity within health systems and a key marker of social justice and health system performance. Although substantial global reductions have been achieved over recent decades, significant disparities persist among population groups, particularly between Indigenous and non-Indigenous populations. These inequities reflect structural differences in timely access to healthcare services, socioeconomic conditions, social determinants of health, and geographic barriers that disproportionately affect Indigenous communities across multiple regions of the world (1–3). Furthermore, they are rooted in longstanding historical processes of marginalization, social exclusion, and the unequal distribution of health resources, which continue to shape health outcomes across generations.

A growing body of evidence has demonstrated that Indigenous women experience a substantially higher risk of maternal mortality than their non-Indigenous counterparts, even within the same national contexts. This excess risk has been linked to multiple interacting factors, including lower coverage and utilization of antenatal care, reduced access to skilled birth attendance and emergency obstetric services, cultural and linguistic barriers, and persistent socioeconomic disadvantage (4–6). These determinants operate within broader structural contexts that influence healthcare access and quality. In addition, territorial dispersion and the limited availability of specialized healthcare services in rural and remote areas contribute to delays in the recognition, referral, and management of obstetric complications, a phenomenon commonly described within the framework of the “three delays” model of maternal mortality (7). Consequently, the unequal geographical distribution of obstetric services constitutes a central dimension of maternal health inequities.

In Latin America, ethnic disparities in maternal health have been extensively documented. Countries such as Bolivia, Guatemala, Peru, and Mexico report significantly higher maternal mortality rates among Indigenous women, highlighting persistent structural inequalities within healthcare systems (8–10). These disparities reflect not only differences in access to healthcare services but also broader social and historical processes associated with structural poverty, social exclusion, discrimination, and the limited incorporation of intercultural approaches into health policies and service delivery (11). As a result, ethnicity continues to be a major determinant of maternal health outcomes throughout the region.

Paraguay faces similar challenges. The country is home to a diverse Indigenous population composed of multiple ethnic groups, many of whom reside in geographically dispersed territories characterized by limited health infrastructure and restricted access to essential healthcare services. National and international reports have consistently shown that Indigenous communities in Paraguay face substantial barriers to accessing adequate healthcare, particularly in the field of maternal health (12,13). These barriers include long travel distances to healthcare facilities, limited availability of emergency obstetric services, socioeconomic inequalities, language barriers, and intercultural challenges that may hinder the effective use of healthcare services. Together, these factors contribute to persistent disparities in maternal health outcomes between Indigenous and non-Indigenous populations.

Moreover, a considerable proportion of maternal deaths are attributable to potentially preventable causes, including obstetric hemorrhage, hypertensive disorders of pregnancy, sepsis, and abortion-related complications. These conditions can be effectively prevented, detected, or treated through timely interventions within well-functioning and equitable health systems (14,15). Therefore, the persistence of such causes not only reflects limitations in the healthcare system’s capacity to provide effective care but also reveals inequities in the territorial distribution of resources, the quality of care provided, and access to comprehensive obstetric services.

Despite the public health importance of this issue, epidemiological evidence examining maternal mortality through a multidimensional framework that integrates territorial, institutional, and intercultural determinants remains limited in Paraguay. In particular, inequalities between Indigenous and non-Indigenous women, as well as disparities across specific Indigenous ethnic groups and territories, remain insufficiently explored in the scientific literature. Addressing these knowledge gaps is essential for informing evidence-based policies aimed at reducing ethnic inequities in maternal health and advancing progress toward national and global commitments to maternal mortality reduction.

Against this background, the aim of this study was to examine structural inequities in maternal mortality in Paraguay between Indigenous and non-Indigenous women by assessing territorial and institutional disparities and differences in the distribution of potentially preventable causes of death through a national population-based analysis.

## Methods

### Study Design

A retrospective analytical observational study was conducted using secondary data. The unit of analysis corresponded to each maternal death recorded during the study period.

### Data Sources

Data were obtained from the national mortality registration system and the official vital statistics corresponding to live births, provided by the General Directorate of Strategic Health Information of the Ministry of Public Health and Social Welfare of Paraguay. The database included nationwide information covering the period from 2014 to 2023.

All maternal deaths registered during the study period were included and defined according to the criteria established by the World Health Organization. The underlying cause of death was coded according to the International Classification of Diseases, Tenth Revision (ICD-10), considering Chapter XV (O00–O99), corresponding to pregnancy, childbirth, and the puerperium, as well as code A34 (obstetric tetanus).

### Study Variables

The primary exposure variable was ethnicity, classified as Indigenous and non-Indigenous.

Ethnicity was conceptualized as a social and structural determinant of health reflecting historical, social, cultural, and territorial inequalities rather than a biological characteristic.

Sociodemographic variables included maternal age, educational level, marital status, specific ethnicity (individual Indigenous groups), department of residence, and department where death occurred. Age was analyzed both as a continuous variable and categorized into age groups.

Institutional variables included the type of institution where death occurred and the area of occurrence (urban or rural). Deaths occurring outside healthcare institutions were defined as those taking place at home, on public roads, or in other locations outside healthcare facilities.

Underlying causes of death were classified as potentially preventable according to the international maternal mortality literature, including obstetric hemorrhage (O72), hypertensive disorders of pregnancy (O14–O15), puerperal sepsis (O85), and abortion-related complications (O03–O06).

### Statistical Analysis

A descriptive and analytical analysis of maternal mortality was performed comparing Indigenous and non-Indigenous women. The maternal mortality ratio (MMR) was calculated as the number of maternal deaths per 100,000 live births and disaggregated by calendar year and ethnic status.

To assess maternal mortality inequities, both absolute and relative measures were estimated. Absolute inequity was expressed as the difference between the maternal mortality ratio among Indigenous women and that among non-Indigenous women, whereas relative inequity was assessed using incidence rate ratios.

The relative concentration of deaths by specific Indigenous ethnic group was evaluated by comparing the observed proportion of deaths with the expected proportion according to their national population share (2.3%), whenever such information was available. A proportion of deaths exceeding the corresponding population share was considered indicative of overrepresentation.

### Temporal Trends

Temporal trends were assessed using Poisson regression models with a log link function. The annual number of maternal deaths was used as the dependent variable, and the logarithm of live births was included as an offset term.

Models included calendar year (continuous variable), ethnic status, and an interaction term between year and ethnicity to evaluate differences in temporal trends between groups. Results were expressed as incidence rate ratios (IRRs) and corresponding 95% confidence intervals (95% CIs).

Additionally, temporal changes in absolute and relative inequities were evaluated using simple linear regression models, with the absolute difference in maternal mortality ratios and the incidence rate ratio as dependent variables, respectively, and calendar year as the independent variable. To facilitate interpretation of regression coefficients, year was centered on the midpoint of the study period.

Given the exceptional increase in maternal mortality observed in 2021, associated with the impact of the COVID-19 pandemic, a sensitivity analysis excluding that year was conducted to assess the robustness of the observed trends.

### Territorial Inequity

To evaluate the territorial distribution of maternal mortality and the potential centralization of obstetric complication management, a Death Centralization Index was calculated for each department. This index was defined as the ratio between the percentage of maternal deaths occurring in a department and the percentage of women residing in that same department.

Values close to 1 indicated balance between place of residence and place of death. Values greater than 1 suggested concentration of deaths within the department, whereas values below 1 indicated displacement of cases to other territories. Since no standardized cut-off points exist for this indicator, a value ≥3 was operationally defined as indicative of extreme centralization.

Additionally, the national proportion of maternal deaths occurring outside the department of residence was estimated as an indicator of structural territorial referral.

### Institutional Inequity

A binary variable for death outside a healthcare institution was constructed and defined as deaths occurring at home, on public roads, or in other non-institutional settings according to the place of death recorded on the death certificate.

The overall proportion of extra-institutional deaths, proportions stratified by ethnicity, and their distribution according to age and educational level were estimated. Crude odds ratios (ORs) and 95% confidence intervals were calculated to assess the association between Indigenous status and death outside a healthcare institution.

Subsequently, a binary logistic regression model was fitted to estimate adjusted odds ratios (aORs), including maternal age (continuous) and educational level (ordinal) as covariates.

### Inequities in Potentially Preventable Causes of Death

Binary variables were created for each specific cause of death and for the overall group of potentially preventable causes. Absolute frequencies and proportions of each cause relative to all maternal deaths were estimated, as well as the overall proportion of potentially preventable maternal deaths.

To evaluate the association between ethnicity and each specific cause, crude odds ratios and corresponding 95% confidence intervals were calculated.

Adjusted binary logistic regression models were subsequently fitted for specific causes that showed evidence of association with Indigenous status in crude analyses. Ethnicity was the primary explanatory variable, while maternal age (continuous) and educational level (ordinal) were included as covariates. Results were reported as adjusted odds ratios (aORs) with corresponding 95% confidence intervals (95% CIs).

### Synthesis of Structural Dimensions of Inequity

Based on the results of the descriptive and analytical analyses, an interpretative synthesis of the structural dimensions of maternal mortality inequities was developed. This synthesis integrated territorial, institutional, and preventability dimensions, allowing the identification of patterns of concentration, exclusion, and structural failures in access to and quality of obstetric care.

Records with incomplete information were excluded only from the corresponding specific analyses. No imputation of missing data was performed.

All analyses were conducted using maximum likelihood-based regression models. Statistical analyses were performed using R software (R Foundation for Statistical Computing, Vienna, Austria).

### Ethical Considerations

The study used anonymized secondary data obtained from national health records and involved no direct contact with participants. The research was conducted in accordance with ethical principles for health research and current national regulations governing the use of secondary data.

## Results

A total of 907 maternal deaths were recorded in Paraguay between 2014 and 2023, of which 112 (12.3%) occurred among Indigenous women and 795 (87.7%) among non-Indigenous women. Considering that Indigenous peoples represent approximately 2.3% of the national population, an overrepresentation ratio of 4.8 was observed, indicating a disproportionate burden of maternal mortality among Indigenous populations (Table 1).

**Table 1.** Sociodemographic and institutional characteristics of maternal deaths according to Indigenous status, Paraguay, 2014–2023.

| Category | Variable | Non-Indigenous n (%) | Indigenous n (%) |
| --- | --- | --- | --- |
| **Age (years)** | <20 | 88 (11.1) | 26 (23.2) |
|  | 20–29 | 272 (34.2) | 53 (47.3) |
|  | 30–39 | 340 (42.8) | 25 (22.3) |
|  | ≥40 | 95 (11.9) | 8 (7.1) |
| **Educational level** | None | 13 (1.6) | 35 (31.2) |
|  | Incomplete primary education | 118 (14.8) | 37 (33.0) |
|  | Complete primary education | 156 (19.6) | 9 (8.0) |
|  | Complete basic education | 83 (10.4) | 7 (6.2) |
|  | Complete secondary education | 111 (14.0) | 0 (0.0) |
|  | Complete high school education | 108 (13.6) | 2 (1.8) |
|  | Higher education | 81 (10.2) | 0 (0.0) |
|  | Unknown | 125 (15.7) | 22 (19.6) |
| **Area of occurrence** | Urban | 641 (80.6) | 89 (79.5) |
|  | Rural | 154 (19.4) | 23 (20.5) |
| **Death outside healthcare institution** | No | 624 (78.5) | 62 (55.4) |
|  | Yes | 171 (21.5) | 50 (44.6) |
| **Total** |  | 795 (87.7) | 112 (12.3) |
| **Overrepresentation ratio\*** |  | 4.8 | — |

\*Calculated as the observed proportion of Indigenous maternal deaths divided by the expected proportion according to the Indigenous population share in Paraguay (2.3%).

Important sociodemographic differences were identified between ethnic groups. Indigenous women exhibited a younger age profile, with 70.5% of deaths occurring among women younger than 30 years, compared with 45.3% among non-Indigenous women. Marked educational inequalities were also observed, as Indigenous women more frequently reported no formal education or incomplete primary education (64.2% versus 16.4%). No substantial differences were identified regarding the area where death occurred (urban or rural). However, deaths occurring outside healthcare institutions were considerably more frequent among Indigenous women than among non-Indigenous women (44.6% versus 21.5%) (Table 1).

Maternal mortality remained consistently higher among Indigenous women throughout the study period (Table 2). The maternal mortality ratio among Indigenous women ranged from 317.7 to 773.6 deaths per 100,000 live births, whereas corresponding values among non-Indigenous women ranged from 58.7 to 145.1 deaths per 100,000 live births. Absolute differences varied between 257.3 and 628.5 deaths per 100,000 live births, while relative measures indicated mortality levels between 4.9 and 8.5 times higher among Indigenous women. The greatest disparity was observed in 2021, when the maternal mortality ratio reached 773.6 among Indigenous women compared with 145.1 among non-Indigenous women.

**Table 2.** Trends in maternal mortality and ethnic inequities among Indigenous and non-Indigenous women in Paraguay, 2014–2023.

| Year | Ethnicity | Maternal deaths n (%) | Live births (n) | Maternal mortality ratio* | Absolute difference† | Incidence rate ratio |
| --- | --- | --- | --- | --- | --- | --- |
| 2014 | Indigenous | 5 (6.9) | 1,574 | 317.7 | 257.3 | 5.3 |
|  | Non-Indigenous | 67 (93.1) | 111,072 | 60.3 | – | – |
| 2015 | Indigenous | 10 (10.5) | 1,844 | 542.3 | 468.0 | 7.3 |
|  | Non-Indigenous | 85 (89.5) | 114,337 | 74.3 | – | – |
| 2016 | Indigenous | 9 (9.4) | 1,725 | 521.7 | 442.2 | 6.6 |
|  | Non-Indigenous | 87 (90.6) | 109,421 | 79.5 | – | – |
| 2017 | Indigenous | 10 (12.8) | 1,967 | 508.4 | 448.7 | 8.5 |
|  | Non-Indigenous | 68 (87.2) | 113,928 | 59.7 | – | – |
| 2018 | Indigenous | 9 (11.4) | 1,98 | 454.5 | 390.7 | 7.1 |
|  | Non-Indigenous | 70 (88.6) | 109,662 | 63.8 | – | – |
| 2019 | Indigenous | 11 (15.1) | 2,22 | 495.5 | 436.8 | 8.4 |
|  | Non-Indigenous | 62 (84.9) | 105,691 | 58.7 | – | – |
| 2020 | Indigenous | 8 (9.9) | 2,236 | 357.8 | 285.1 | 4.9 |
|  | Non-Indigenous | 73 (90.1) | 100,486 | 72.6 | – | – |
| 2021 | Indigenous | 19 (11.4) | 2,456 | 773.6 | 628.5 | 5.3 |
|  | Non-Indigenous | 147 (88.6) | 101,31 | 145.1 | – | – |
| 2022 | Indigenous | 13 (14.8) | 2,875 | 452.2 | 373.6 | 5.8 |
|  | Non-Indigenous | 75 (85.2) | 95,43 | 78.6 | – | – |
| 2023 | Indigenous | 18 (22.8) | 3,313 | 543.3 | 474.8 | 7.9 |
|  | Non-Indigenous | 61 (77.2) | 89,086 | 68.5 | – | – |

\*Maternal mortality ratio per 100,000 live births.
†Difference between Indigenous and non-Indigenous maternal mortality ratios.

Temporal trend analyses demonstrated persistently high absolute inequalities in maternal mortality across the study period. The annual absolute difference showed an average increase of 10.8 maternal deaths per 100,000 live births. In a sensitivity analysis excluding 2021, a year characterized by the COVID-19 pandemic, the trend remained positive, with an average increase of 4.2 maternal deaths per 100,000 live births per year. These findings indicate that ethnic inequities in maternal mortality persisted beyond the pandemic context (Figure 1).

**Figure 1.** Temporal trends in absolute maternal mortality inequities between Indigenous and non-Indigenous women in Paraguay, 2014–2023. Note. Temporal trends in absolute maternal mortality inequities between Indigenous and non-Indigenous women in Paraguay, 2014–2023. The solid line represents the annual absolute difference in maternal mortality ratios (maternal deaths per 100,000 live births) between Indigenous and non-Indigenous women. The dashed line represents the fitted linear trend.

In the Poisson regression model adjusted for the number of live births, maternal mortality increased by an average of 3.9% annually (incidence rate ratio [IRR] = 1.04; 95% confidence interval [95% CI]: 1.01–1.07; p = 0.002). The interaction between calendar year and Indigenous status was not statistically significant (IRR = 0.99; 95% CI: 0.95–1.03; p = 0.680), suggesting that temporal trends evolved similarly among Indigenous and non-Indigenous women (Supplementary Table 1).

Overall, 220 maternal deaths (24.3%) occurred outside healthcare institutions. This proportion was substantially higher among Indigenous women than among non-Indigenous women (44.6% versus 21.5%) (Table 3).

**Table 3.** Maternal deaths occurring outside healthcare institutions according to Indigenous status, Paraguay, 2014–2023.

| **Ethnicity** | **Death outside**<br>**healthcare institution**<br>**n (%)** | **Death within**<br>**healthcare institution**<br>**n (%)** | **Total (n)** |
| --- | --- | --- | --- |
| Non-Indigenous | 171 (21.5) | 624 (78.5) | 795 |
| Indigenous | 50 (44.6) | 62 (55.4) | 112 |
| **Total** | **220 (24.3)** | **686 (75.7)** | **907** |

Note. Deaths outside healthcare institutions were defined as deaths occurring at home, on public roads, or in other non-institutional settings.

Indigenous status was strongly associated with maternal deaths occurring outside healthcare institutions. In crude analyses, Indigenous women had nearly three times greater odds of dying outside a healthcare facility than non-Indigenous women (OR = 2.94; 95% CI: 1.94–4.46; p < 0.001). After adjustment for maternal age and educational level, the association remained strong and statistically significant (aOR = 3.41; 95% CI: 2.20–5.29; p < 0.001). Maternal age was positively associated with the outcome (aOR = 1.02 per year; 95% CI: 1.00–1.04; p = 0.030), whereas educational level was not significantly associated (aOR = 1.03; 95% CI: 0.96–1.10; p = 0.470) (Table 4; Supplementary Table 2).

**Table 4.** Association between Indigenous status and maternal deaths occurring outside healthcare institutions, Paraguay, 2014–2023.

| Variable | Crude OR (95% CI) | p-value | Adjusted OR (95% CI)* | p-value |
| --- | --- | --- | --- | --- |
| Indigenous ethnicity | 2.94 (1.94–4.46) | <0.001 | 3.41 (2.20–5.29) | <0.001 |
| Maternal age (per year) | – | – | 1.02 (1.00–1.04) | 0.030 |
| Educational level | – | – | NS | >0.05 |

\*Adjusted for maternal age and educational level.
OR = odds ratio; CI = confidence interval; NS = not statistically significant.

Potentially preventable causes accounted for 386 maternal deaths (42.4% of all maternal deaths). Among deaths occurring outside healthcare institutions, 42.1% were attributed to preventable causes, indicating a substantial burden of potentially avoidable mortality.

Among the specific causes analyzed, obstetric hemorrhage accounted for 12.75% of all maternal deaths and showed a strong association with Indigenous status. Indigenous women had significantly greater odds of dying from obstetric hemorrhage both in crude analyses (OR = 3.79; 95% CI: 2.39–6.02; p < 0.001) and after adjustment for maternal age and educational level (aOR = 3.83; 95% CI: 2.31–6.37; p < 0.001). Puerperal sepsis was also associated with Indigenous status in crude analyses (OR = 2.81; 95% CI: 1.27–6.20; p = 0.011), although statistical significance was lost after adjustment (aOR = 2.20; 95% CI: 0.95–5.10; p = 0.060). No significant associations were identified for hypertensive disorders of pregnancy or abortion-related complications (Table 5).

**Table 5.** Association between Indigenous status and potentially preventable causes of maternal death, Paraguay, 2014–2023.

| **Cause (ICD-10)** | **n (%) of all maternal deaths** | **Crude OR** | **95% CI** | **p-value** | **Adjusted OR\*** | **95% CI** | **p-value** |
| --- | --- | --- | --- | --- | --- | --- | --- |
| Obstetric hemorrhage (O72) | 116 (12.75) | 3.79 | 2.39–6.02 | <0.001 | 3.83 | 2.31–6.37 | <0.001 |
| Hypertensive disorders of pregnancy (O14–O15) | 142 (15.60) | 1.29 | 0.77–2.15 | 0.337 | – | – | – |
| Puerperal sepsis (O85) | 33 (3.63) | 2.81 | 1.27–6.20 | 0.011 | 2.20 | 0.95–5.10 | 0.060 |
| Abortion-related complications (O03–O06) | 95 (10.44) | 0.54 | 0.24–1.19 | 0.124 | – | – | – |
| **Total potentially preventable causes** | **386 (42.40)** | – | – | – | – | – | – |

\*Adjusted models were fitted for causes showing evidence of association in crude analyses.
OR = odds ratio; CI = confidence interval.

Taken together, these findings reveal a persistent pattern of structural inequities in maternal mortality in Paraguay. Indigenous women were disproportionately affected by maternal mortality, experienced consistently higher mortality ratios throughout the study period, were significantly more likely to die outside healthcare institutions, and faced a substantial burden of potentially preventable causes of death. These findings suggest the coexistence of territorial, institutional, and healthcare access barriers that disproportionately affect Indigenous populations and contribute to enduring ethnic inequities in maternal health outcomes (Supplementary Figure 2).

## Discussion

The findings of the present study reveal profound structural inequities in maternal mortality in Paraguay, consistently manifested across territorial, institutional, and preventability dimensions. Taken together, these results suggest that maternal mortality is not solely the consequence of individual-level factors but also reflects systemic failures related to the organization, accessibility, responsiveness, and quality of healthcare services. Furthermore, the persistence of these disparities over an entire decade indicates that they represent deeply rooted structural phenomena rather than temporary or circumstantial events. From an equity perspective, these findings highlight the enduring influence of social, territorial, and ethnic determinants on maternal health outcomes. Although this study focuses on Paraguay, the observed patterns are consistent with evidence from Indigenous populations in other settings, where maternal mortality continues to be shaped by structural inequities, geographic marginalization, and unequal access to quality healthcare services.

From a territorial perspective, the marked concentration of maternal deaths in specific departments (Death Centralization Index = 5.74), together with the identification of territories functioning as “exporters” of patients, reflects an unequal organization of the healthcare system in which specialized services remain concentrated in a limited number of centers. Similar patterns have been described in other Latin American settings, where fragmented healthcare networks contribute to delays in referral processes and increase the risk of preventable maternal mortality (16,17). The high proportion of deaths occurring outside the department of residence further supports the existence of structural dependence on centralized healthcare networks, potentially compromising timely access to emergency obstetric care. These findings can also be interpreted within the framework of the three delays model, particularly regarding delays in reaching appropriate healthcare facilities, a situation frequently observed in geographically dispersed settings with limited availability of specialized services.

At the institutional level, 24.3% of maternal deaths occurred outside healthcare institutions, a proportion that reached 44.6% among Indigenous women, revealing substantial inequalities in access to formal healthcare services. This disparity remained significant after adjustment for age and educational level (adjusted odds ratio = 3.41; 95% confidence interval: 2.20–5.29), indicating that Indigenous status constitutes an independent determinant of exclusion from institutional maternal healthcare. These findings are consistent with previous studies documenting geographic, economic, cultural, and linguistic barriers that limit access to maternal healthcare among Indigenous populations (5,18). In this context, the high frequency of extra-institutional deaths may be interpreted as a critical indicator of deficiencies in healthcare accessibility, acceptability, and continuity of care. Moreover, this pattern suggests the persistence of inequalities associated with both the first and second delays of the maternal mortality framework.

Regarding preventability, the finding that 42.4% of maternal deaths were attributed to preventable causes, together with a similar proportion of preventable causes among deaths occurring outside the healthcare system (42.1%), indicates a substantial burden of potentially avoidable mortality. These results suggest that the observed limitations extend beyond access barriers and also involve deficiencies in the quality and effectiveness of care delivered within the healthcare system itself. Obstetric hemorrhage and puerperal sepsis, both largely preventable through timely diagnosis and treatment, showed relevant associations with Indigenous status, reinforcing the existence of inequities in access to and delivery of emergency obstetric care (19,20). The absence of substantial differences in the proportion of preventable causes between deaths occurring inside and outside healthcare institutions suggests a cross-cutting problem in which limitations in healthcare coverage coexist with deficiencies in the capacity of services to provide timely and effective care. These findings are also consistent with the third delay of the maternal mortality model, related to delays in receiving appropriate care after contact with healthcare services.

An additional finding of particular relevance was the overrepresentation ratio of 4.8, indicating that Indigenous women experienced a burden of maternal mortality nearly five times higher than expected according to their proportion within the national population. This represents robust evidence of structural inequity and is consistent with the social determinants of health framework, which recognizes the influence of historical, social, economic, and political factors on the unequal distribution of health outcomes (21). The magnitude of this overrepresentation suggests that improvements achieved in maternal health at the national level have not benefited all population groups equally, leaving Indigenous women disproportionately exposed to preventable risks and adverse maternal outcomes. These disparities likely reflect the intersection of ethnicity, educational disadvantage, territorial marginalization, and barriers to healthcare access, highlighting the cumulative and mutually reinforcing nature of maternal health inequities.

Another important finding was the persistence of inequalities throughout the study period. Despite annual fluctuations, both absolute and relative disparities between Indigenous and non-Indigenous women remained substantial over the decade analyzed. Even after excluding 2021, a year heavily affected by the COVID-19 pandemic, positive trends persisted, indicating that the observed inequities cannot be attributed solely to the pandemic context. Instead, these findings point to the presence of enduring structural mechanisms that perpetuate maternal health inequalities and hinder sustainable reductions in ethnic disparities. The persistence of these gaps over time suggests that improvements in maternal health indicators alone may be insufficient unless accompanied by policies specifically designed to address the underlying social and structural determinants of inequity.

Several limitations should be acknowledged. First, the use of secondary mortality data may be subject to misclassification, underreporting, and variability in data quality, particularly for variables such as ethnicity and educational level. Second, the observational nature of the study limits causal inference, and the associations identified should therefore be interpreted cautiously. Third, some estimates, particularly those related to specific causes such as puerperal sepsis, may have been influenced by the limited number of events, resulting in wider confidence intervals and reduced precision of adjusted estimates. Finally, it was not possible to incorporate other potentially relevant variables, including socioeconomic conditions, geographic accessibility to healthcare services, or contextual territorial characteristics, which could have provided a more comprehensive understanding of the inequities identified.

Overall, these findings demonstrate that maternal mortality in Paraguay remains a persistent problem of structural inequity characterized by ethnic, territorial, and institutional disparities that disproportionately affect Indigenous women. The territorial concentration of deaths, the higher frequency of deaths occurring outside healthcare institutions, the substantial burden of potentially preventable causes, and the persistence of inequalities over time collectively reveal structural barriers affecting both access to and quality of obstetric care. These findings underscore the need for comprehensive equity-oriented strategies, including the decentralization of healthcare services, strengthening of obstetric care networks, implementation of intercultural approaches to maternal healthcare, and enhancement of the healthcare system’s capacity to provide timely and effective obstetric care. Addressing maternal mortality among Indigenous women therefore requires not only improvements in healthcare delivery but also sustained efforts to reduce the broader social and structural inequities that shape maternal health outcomes. Such measures are essential to reduce preventable maternal mortality and advance maternal health equity among Indigenous populations.

## Data Availability

The data used in this study were obtained from the Directorate General of Health Information (Dirección General de Información Estratégica en Salud, DIGIES) of the Ministry of Public Health and Social Welfare of Paraguay. Due to legal and ethical restrictions governing the use of individual-level mortality data, the datasets are not publicly available. Researchers may request access to the data directly from the Ministry of Public Health and Social Welfare of Paraguay, subject to institutional approval and applicable regulations. All aggregated data supporting the findings of this study are included within the manuscript and its Supporting Information files.

## DECLARATIONS

### Ethics approval and consent to participate

Not applicable.

### Consent for publication

Not applicable.

### Author contributions

Conceptualization: DDZ and PR; Data curation: DDZ and JETG; Formal analysis: DDZ, and JETG; Investigation: DDZ, JETG and PR; Methodology: DDZ and PR; Supervision: DDZ, JETG and PR; Writing original draft: DDZ, BN and PR; Writing review and editing: DDZ, JETG, BN and PR. All authors read and approved the final manuscript.

## Acknowledgements

The authors gratefully acknowledge the General Directorate of Strategic Health Information (DIGIES), Ministry of Public Health and Social Welfare (MSPYBS), Asunción, Paraguay, for their commitment to strengthening health information systems and for providing access to the data that made this study possible.

## Data Availability Statement

The data underlying this study were obtained from the Ministry of Public Health and Social Welfare of Paraguay through the General Directorate of Strategic Health Information (DIGIES). Due to institutional and administrative restrictions, the data are not publicly available. Access to the data may be granted upon reasonable request and with prior authorization from the Ministry of Public Health and Social Welfare of Paraguay.

## SUPPLEMENTARY MATERIAL

Supplementary Table 1.

Supplementary Figure 1.

Supplementary Figure 2.

